# Coverage and Effectiveness of mRNA COVID-19 Vaccines among Veterans

**DOI:** 10.1101/2021.06.14.21258906

**Authors:** Yinong Young-Xu, Caroline Korves, Jeff Roberts, Ethan I. Powell, Gabrielle M. Zwain, Jeremy Smith, Hector S. Izurieta

## Abstract

**Importance:** The effectiveness of mRNA vaccination in a large and diverse American population, with older age and higher co-morbidity has not been assessed.

**Objective:** To describe the scope of the mRNA vaccination rollout among the diverse U.S. Veterans population, and to study the mRNA COVID-19 vaccine effectiveness (VE) against infection, symptomatic disease, hospitalization, and death.

**Methods:** Vaccination histories were obtained from medical records to determine if patients tested for SARS-CoV-2 were unvaccinated, partially vaccinated (first dose of mRNA COVID-19 vaccine), or fully vaccinated (two doses) at time of testing. First, coverage with any COVID-19 vaccination was described for all Veterans enrolled in Veterans Health Administration (VHA). Second, to evaluate VE, a matched test-negative case-control evaluation was conducted utilizing SARS-CoV-2 positive (cases [n=16,690]) and SARS-CoV-2 negative (controls [n=61,610]) tests from Veterans aged ≥18 years old who routinely sought care at a VHA facility and were tested from December 14, 2020, through March 14, 2021. VE was calculated from odds ratios (ORs) with 95% confidence intervals (CI).

**Results:** By March 7, 2021, among 6,170,750 Veterans, 1,547,045 (23%) received at least one COVID-19 vaccination. mRNA COVID-19 VE against infection, regardless of symptoms, was 94% (95% CI 92-95) and 58% (95%CI 54-62) for full and partial vaccination (vs. no vaccination), respectively. VE against infection was similar across subpopulations, and it was not significantly different from VE against symptomatic disease. VE against COVID-19-related hospitalization and death for full vs. no vaccination was 89% (95%CI 81-93) and 99% (95%CI 87-100), respectively.

**Conclusions and Relevance:** The VHA’s efficient and equitable distribution of effective vaccines decreased COVID-19 infections, hospitalization, and mortality similarly for all Veterans, including Veterans with low income, homeless Veterans, immunocompromised, the elderly, minorities, and rural Veterans thus reducing health inequalities.

## INTRODUCTION

On December 11, 2020, the United States (U.S.) Food and Drug Administration (FDA) issued an Emergency Use Authorization (EUA) for Pfizer-BioNTech COVID-19 Vaccine, for the prevention of coronavirus disease 2019 (COVID-19) for individuals 16 years and older(1). One week later, the FDA issued an EUA for Moderna COVID-19 Vaccine, developed by Moderna, for individuals 18 years and older(2); both vaccines are mRNA vaccines. Prior to these EUAs, the pandemic’s impact on Veterans enrolled in the Veterans Health Administration (VHA), like elsewhere in the US, was devastating. Approximately 207,000 COVID-19 cases had been reported by the end of 2020 among VHA enrolled Veterans(3). Additionally, the burden of COVID-19 cases was distributed disproportionally among racial and ethnic minorities and could not be explained by underlying chronic conditions. About 35% and 13% of infected Veterans were Black and Hispanic, although they comprised only 13% and 7% of the total VHA patient population, respectively. Over 10,000 of these patients died(3).

The VHA has worked closely with the Centers for Disease Control and Prevention (CDC) and other federal partners to provide COVID-19 vaccines to Veterans as quickly and safely as possible. On March 24, 2021 the success of the VHA vaccination program prompted the U.S. Congress to pass the “Strengthening and Amplifying Vaccination Efforts to Locally Immunize All Veterans and Every Spouse Act” which authorized the Department of Veterans Affairs (VA) to expand its vaccination effort beyond enrolled Veterans(4). Rapid deployment of the vaccination program was accompanied by VHA’s nationwide SARS-CoV-2 testing effort, which aimed to test and record both symptomatic and asymptomatic patients across all VHA facilities. Combined with accurate and timely recording of vaccination, this created an opportunity for a robust and well-powered test-negative case-control analysis.

Aiming to describe the extent of vaccination coverage and evaluate the effectiveness of both mRNA vaccines in a diverse population that included socio-economically disadvantaged and medically high-risk individuals, we focused this study on the first three months of the vaccination effort in the VHA, thus the analysis excluded data on the Janssen COVID-19 Vaccine.

## METHODS

The study protocol was approved by the institutional review board of the VA Medical Center in White River Junction, VT.

### Data Source

The VHA is the largest integrated health care system in the U.S., providing comprehensive care to over nine million Veterans at more than 171 medical centers and 1,112 outpatient sites of care(4). We analyzed electronic medical record data from the VHA Corporate Data Warehouse (CDW). The CDW contains patient-level information on all patient encounters, treatments, prescriptions (including vaccinations), and laboratory results rendered in VHA medical facilities.

### Study Design

First, vaccination coverage, defined as having at least one COVID-19 vaccination administered at a VHA facility between December 14, 2020 and March 7, 2021, was described for the population of VHA enrollees. Second, we conducted a test-negative design (TND) case-control study to evaluate mRNA COVID-19 VE against infection, irrespective of symptoms, and a case-control study to evaluate VE against COVID-19-related hospitalization and death. The study population included Veterans ages 18 years and older, with residence in a U.S. state or Washington, D.C., who presented for SARS-CoV-2 PCR or antigen testing at a VHA outpatient or emergency room facility, or had testing within one day of hospitalization, during the study period (December 14, 2020, and March 7, 2021). Patients were required to have had VHA enrollment for at least two years prior to the study period and at least one inpatient or two outpatient visits in the past two years. Individuals meeting any of the following criteria were excluded: a COVID-19diagnosis and/or a positive SARS-CoV-2 antigen or PCR test at any time between February 2020 and study initiation (December 14, 2020); hospitalization more than one day prior to testing; incomplete VHA medical records.

For the TND case-control study to assess VE against infection, positive SARS-CoV-2 tests from qualifying patients were classified as cases. Negative SARS-CoV-2 tests from qualifying patients served as controls, and a maximum of four controls were matched to each case based on Health and Human Services (HHS) geographic region, and testing date (within 14 days of case testing date) as both of these factors are related to local disease burden, likelihood of testing positive for SARS-CoV-2, and vaccine exposure status. A TND case-control study to assess VE against symptomatic disease was also conducted. Within the population of patients with a SARS-CoV-2 test performed, a case-control study was also conducted to assess VE in preventing COVID-19-related hospitalization and death. For these analyses, cases were those who tested positive and were hospitalized (or died within 30 days of testing positive), and controls were Veterans who were tested negative and did not have the outcome of interest. Up to four controls were matched to each case based on geographic region and testing date.

### Exposure, Outcome and Covariate Assessment

Vaccination status was based on records of mRNA COVID-19 vaccination at a VHA facility from December 14, 2020-February 28, 2021. A person was classified as unvaccinated until the day prior to the first vaccination, partially vaccinated from day 7 after the first vaccination until the day prior to the second vaccination, and fully vaccinated starting at 7 days after the second vaccination (*Supplemental* Figure 1). Days 0-6 after the first and second doses were excluded from all analyses. Vaccination status was determined at the date of testing. For the TND case-control study to assess VE against infection, medical records were used to identify cases (positive tests) and controls (negative tests). For the TND case-control study to assess VE against symptomatic disease, Veterans who tested positive were further restricted to those who, on the day of testing, had evidence of at least one COVID-19 symptom (*Supplemental* Table 1). For the case-control study for hospitalization and death, a COVID-19-related hospitalization was identified by the presence of an admission and discharge diagnosis of COVID-19 occurring any time after the first positive SARS-CoV-2 test. A death occurring in hospital with a COVID-19 discharge diagnosis or a death occurring within 30 days of a positive SARS-CoV-2 test was classified as a COVID-19-related death; controls were drawn from patients with a negative SARS-CoV-2 test who were not hospitalized for COVID-19 during the study period (or who did not die within 30 days of SARS-CoV-2 test).

Demographic and clinical characteristics of patients tested for SARS-CoV-2 were assessed at the time of testing or based on data from the prior two-year period, for characteristics such as comorbidities, Charlson Comorbidity Index and Care Assessment Needs (CAN) score (See *Supplemental* Table 1 for full definitions).

### Statistical Analyses

For VHA enrollees and for subpopulations, vaccination coverage was reported as the frequency and proportion of individuals with at least one vaccination.

For the TND case-control study, for cases (positive SARS-CoV-2 test) and controls (negative SARS-CoV-2 test), baseline demographic and clinical characteristics of the tested patients were described by reporting frequency and proportion for categorical variables and median interquartile range (IQR) for continuous variables. Missing data were reported. Standardized mean difference (SMD) was used to describe differences in characteristics between cases and controls. We used conditional logistic regression to calculate odds ratios (ORs) with 95% confidence interval (CI) for the association between positive SARS-CoV-2 testing and receipt of mRNA COVID-19 vaccine. VE was calculated as (1− OR_odds of SARS-CoV-2 in vaccinated vs odds of SARS-CoV-2 in unvaccinated_) x 100%. Analyses were conducted comparing partial vaccination, days 14-20 after first dose, full vaccination, and late full vaccination (day 14 after second dose and onwards) versus no vaccination. Models for adjusted analyses included covariates for potential confounders of the association between vaccination status and testing positive for SARS-CoV-2. Conditional logistic regression for the case-control analyses of hospitalization and death and for VE against symptomatic disease were conducted similarly.

All analyses were conducted using SAS, version 7.

## RESULTS

### Vaccination Coverage

By March 7, 2021, 1,547,045 (23.1%) VHA enrollees were administered at least one dose of a COVID-19 vaccine at a VHA facility (Table 1). Vaccination coverage was higher (33.8%) among Veterans ≥65 years old. Vaccination coverage for this early vaccination period was 20.6%, 24.9%, and 23.9% for Hispanics, Blacks and Whites, respectively. Vaccination reached homeless Veterans (19.3%), those living in nursing homes (40.9%) (Table 1).

**Table 1.** COVID-19 Vaccination Coverage through March 7, 2021 among VHA Enrollees^a^

|  | Number of enrollees<br>[A] | Vaccinated enrollees<br>[B] | % Vaccination coverage<br>[B/A] |
| --- | --- | --- | --- |
| All enrollees | 6,710,750 | 1,547,045 | 23% |
| Age range, y |  |  |  |
| 18-44 | 1,382,988 | 63,794 | 5% |
| 45-64 | 1,950,293 | 340,959 | 18% |
| 65-74 | 1,895,735 | 625,177 | 33% |
| 75-84 | 1,000,503 | 367,395 | 37% |
| ≥85 | 481,231 | 149,720 | 31% |
| ≥65 | 3,377,469 | 1,142,292 | 34% |
| Sex |  |  |  |
| Female | 634,302 | 95,579 | 15% |
| Male | 6,076,448 | 1,451,466 | 24% |
| Race or ethnic group |  |  |  |
| Black | 1,110,223 | 276,750 | 25% |
| Hispanic | 464,284 | 95,845 | 21% |
| White | 4,407,279 | 1,051,749 | 24% |
| Other | 384,981 | 80,993 | 21% |
| Missing | 343,983 | 41,708 | 12% |
| Urban-rural classification |  |  |  |
| Highly rural | 83,386 | 13,649 | 16% |
| Rural | 2,176,409 | 437,793 | 20% |
| Urban | 4,450,955 | 1,095,603 | 25% |
| VHA defined priority group |  |  |  |
| 1-4 | 2,545,311 | 552,638 | 22% |
| 5-6 | 2,156,949 | 475,988 | 22% |
| 7-8 | 2,008,490 | 518,419 | 26% |
| Nursing home resident | 32,063 | 13,103 | 41% |
| Homeless | 124,850 | 24,103 | 19% |
| Low Income | 663,649 | 138,463 | 21% |
| Comorbidities |  |  |  |
| Asthma | 336,849 | 135,672 | 40% |
| Cancer | 191,185 | 91,660 | 48% |
| Cancer, metastatic | 20,768 | 10,520 | 51% |
| Coronary artery disease | 367,556 | 161,922 | 44% |
| Congestive heart failure | 166,994 | 76,177 | 46% |
| Chronic heart disease | 212,324 | 97,836 | 46% |
| Chronic obstructive pulmonary disease | 363,203 | 146,639 | 40% |
| Cardiovascular disease | 114,495 | 48,279 | 42% |
| Dementia | 72,101 | 26,014 | 36% |
| Dyslipidemia | 745,217 | 289,270 | 39% |
| Human immunodeficiency virus (HIV) | 19,422 | 7,758 | 40% |
| Hypertension | 1,473,558 | 572,278 | 39% |
| Liver disease, mild | 94,328 | 36,314 | 38% |
| Liver disease, severe | 8,066 | 3,336 | 41% |
| Myocardial infarction | 36,544 | 16,591 | 45% |
| Hemiplegia or paraplegia | 19,979 | 9,048 | 45% |
| Peptic ulcer disease | 8,683 | 3,761 | 43% |
| Peripheral vascular disease | 140,128 | 65,357 | 47% |
| Rheumatoid arthritis | 43,543 | 17,674 | 41% |
| Renal disease | 219,051 | 100,879 | 46% |
| Immunocompromised | 455,601 | 206,098 | 45% |
| Diabetes mellitus | 967,034 | 384,596 | 40% |
| <sup>a</sup>Definitions for urban-rural classification, VHA defined priority group, and comorbidities are provided in Supplemental Table 1. |  |  |  |

### Study Population

We identified 15,404 positive SARS-CoV-2 tests, classified as cases, and 497,584 negative tests, classified as controls (Figure 1). After matching, there were 15,404 cases and 61,610 controls. Baseline characteristics for cases and controls before and after matching are shown in Table 2. At the time of testing, 14,799 (96%) cases and 50,831 (83%) controls were unvaccinated in the matched analysis. Compared with controls, cases were younger and more likely to be White.

**Table 2.** Exposure Status and Baseline Characteristics of Study Participants^a^

|  | Unmatched |  |  | Matched |  |  |
| --- | --- | --- | --- | --- | --- | --- |
|  | Cases<br>(n=15,404) | Controls<br>(n=497,584) | SMD<sup>b</sup> | Cases<br>(n=15,404) | Controls<br>(n=61,610) | SMD<sup>b</sup> |
| Vaccination status, No. (%) |  |  |  |  |  |  |
| Full | 72 (1) | 34,979 (7) | 35 | 72 (1) | 4,959 (8) | 38 |
| Partial | 533 (4) | 36,545 (7) | 17 | 533 (4) | 5,820 (9) | 25 |
| None | 14,799 (96) | 426,060 (86) | 37 | 14,799 (96) | 50,831 (83) | 45 |
| Vaccine manufacturer, No. (%) |  |  |  |  |  |  |
| Moderna | 342 (2) | 39,007 (8) | 26 | 342 (2) | 6,182 (10) | 33 |
| Pfizer-BioNTech | 263 (2) | 32,517 (7) | 24 | 263 (2) | 4,597 (8) | 28 |
| None<br>(unvaccinated) | 14,799 (96) | 426,060 (86) | 37 | 14,799 (96) | 50,831 (83) | 45 |
| Month of Test, No. (%) |  |  |  |  |  |  |
| 2020-Dec. | 4,603 (30) | 111,052 (22) | 17 | 4,603 (30) | 18,349 (30) | 0 |
| 2021-Jan. | 7,306 (47) | 190,353 (38) | 18 | 7,306 (47) | 25,504 (41) | 12 |
| 2021-Feb. | 2,962 (19) | 155,601 (31) | 28 | 2,962 (19) | 14,537 (24) | 11 |
| 2021-Mar. | 533 (4) | 40,578 (8) | 20 | 533 (4) | 3,220 (5) | 9 |
| Care setting for test, No. (%) |  |  |  |  |  |  |
| ED/At Admission | 6,959 (45) | 119,201 (24) | 46 | 6,959 (45) | 15,982 (26) | 41 |
| Outpatient | 8,445 (55) | 378,383 (76) | 46 | 8,445 (55) | 45,629 (74) | 41 |
| Age, years |  |  |  |  |  |  |
| 18-44 | 2,535 (17) | 73,236 (15) | 5 | 2,535 (17) | 668 (1) | 56 |
| 45-64 | 5,409 (35) | 177,629 (36) | 1 | 5,409 (35) | 22,642 (37) | 3 |
| 65-74 | 4,757 (31) | 160,157 (32) | 3 | 4,757 (31) | 24,673 (40) | 19 |
| 75-84 | 1,928 (13) | 64,096 (13) | 1 | 1,928 (13) | 9,481 (15) | 8 |
| 85+ | 775 (5) | 22,466 (5) | 2 | 775 (5) | 4,147 (7) | 7 |
| Sex, No. (%) |  |  |  |  |  |  |
| Female | 1,457 (10) | 55,591 (11) | 6 | 1,457 (10) | 5,868 (10) | 0 |
| Male | 13,947 (91) | 441,993 (89) | 6 | 13,947 (91) | 55,742 (91) | 0 |
| Race, No. (%) |  |  |  |  |  |  |
| Black | 3,848 (25) | 121,718 (25) | 1 | 3,848 (25) | 19,807 (32) | 16 |
| Hispanic | 1,014 (7) | 34,816 (7) | 2 | 1,014 (7) | 2,884 (5) | 8 |
| White | 9,620 (63) | 303,234 (61) | 3 | 9,620 (63) | 34,731 (56) | 12 |
| Other | 922 (6) | 37,816 (8) | 6 | 922 (6) | 4,188 (7) | 3 |
| Rurality, No. (%) |  |  |  |  |  |  |
| Highly rural | 107 (1) | 3,015 (1) | 1 | 107 (1) | 376 (1) | 1 |
| Rural | 4,667 (30) | 120,555 (24) | 14 | 4,667 (30) | 14,758 (24) | 14 |
| Urban | 10,630 (69) | 374,014 (75) | 14 | 10,630 (69) | 46,476 (75) | 14 |
| HHS region, No. (%) |  |  |  |  |  |  |
| Region 1 | 803 (5) | 23,289 (5) | 2 | 803 (5) | 3,212 (5) | 0 |
| Region 2 | 763 (5) | 23,723 (5) | 1 | 763 (5) | 3,052 (5) | 0 |
| Region 3 | 2,286 (15) | 48,958 (10) | 15 | 2,286 (15) | 9,143 (15) | 0 |
| Region 4 | 3,944 (26) | 138,087 (28) | 4 | 3,944 (26) | 15,976 (26) | 0 |
| Region 5 | 1,661 (11) | 67,333 (14) | 8 | 1,661 (11) | 6,644 (11) | 0 |
| Region 6 | 3,425 (22) | 58,556 (12) | 28 | 3,425 (22) | 13,696 (22) | 0 |
| Region 7 | 714 (5) | 23,441 (5) | 0 | 714 (5) | 2,856 (5) | 0 |
| Region 8 | 445 (3) | 15,880 (3) | 2 | 445 (3) | 1,780 (3) | 0 |
| Region 9 | 976 (6) | 80,345 (16) | 31 | 976 (6) | 3,904 (6) | 0 |
| Region 10 | 337 (2) | 17,946 (4) | 8 | 337 (2) | 1,348 (2) | 0 |
| Homeless, No. (%) | 620 (4) | 32,933 (7) | 12 | 620 (4) | 4,646 (8) | 15 |
| Low income, No. (%) | 2,139 (14) | 77,418 (16) | 5 | 2,139 (14) | 12,059 (20) | 15 |
| Nursing home use, No. (%) | 124 (1) | 9,212 (2) | 9 | 124 (1) | 2,021 (3) | 18 |
| VA priority group, No. (%) |  |  |  |  |  |  |
| 1-4 | 5,823 (38) | 210,310 (42) | 9 | 5,823 (38) | 32,276 (52) | 29 |
| 5-6 | 5,131 (33) | 161,290 (32) | 2 | 5,131 (33) | 17,182 (28) | 12 |
| 7-8 | 4,450 (29) | 125,984 (25) | 8 | 4,450 (29) | 12,152 (20) | 22 |
| Quan's CCI |  |  |  |  |  |  |
| Median (IQR) | 0 (0, 2) | 1 (0, 2) |  | 0 (0, 2) | 1 (0, 3) |  |
| BMI, No. (%) |  |  |  |  |  |  |
| Overweight/ obese | 7,654 (50) | 226,955 (46) | 8 | 7,654 (50) | 26,933 (44) | 12 |
| Comorbidities |  |  |  |  |  |  |
| Asthma | 1,726 (11) | 68,761 (14) | 8 | 1,726 (11) | 11,803 (19) | 22 |
| Cancer | 992 (6) | 40,710 (8) | 7 | 992 (6) | 5,923 (10) | 12 |
| Cancer (metastatic) | 148 (1) | 7,718 (2) | 5 | 148 (1) | 1,181 (2) | 8 |
| Coronary artery disease | 2,018 (13) | 68,512 (14) | 2 | 2,018 (13) | 11,050 (18) | 13 |
| Congestive heart failure | 1,208 (8) | 45,941 (9) | 5 | 1,208 (8) | 7,857 (13) | 16 |
| Chronic kidney disease | 1,458 (10) | 50,718 (10) | 2 | 1,458 (10) | 8,585 (14) | 14 |
| Chronic obstructive pulmonary disease | 1,866 (12) | 74,799 (15) | 9 | 1,866 (12) | 12,812 (21) | 24 |
| Cardiovascular disease | 676 (4) | 30,431 (6) | 8 | 676 (4) | 5,464 (9) | 18 |
| Dementia | 489 (3) | 30,981 (6) | 14 | 489 (3) | 6,619 (11) | 30 |
| Diabetes mellitus (complicated) | 1,827 (12) | 58,907 (12) | 0 | 1,827 (12) | 10,107 (16) | 13 |
| Diabetes mellitus (uncomplicated) | 2,830 (18) | 77,950 (16) | 7 | 2,830 (18) | 11,647 (19) | 1 |
| Dyslipidemia | 3,574 (23) | 122,001 (25) | 3 | 3,574 (23) | 18,732 (30) | 16 |
| HIV | 82 (1) | 4,069 (1) | 3 | 82 (1) | 923 (2) | 10 |
| Hypertension | 6,694 (44) | 217,124 (44) | 0 | 6,694 (44) | 33,962 (55) | 23 |
| Liver disease (mild) | 578 (4) | 24,199 (5) | 5 | 578 (4) | 4,098 (7) | 13 |
| Liver disease (moderate/severe) | 78 (1) | 3,781 (1) | 3 | 78 (1) | 584 (1) | 5 |
| Myocardial infarction (history) | 334 (2) | 13,182 (3) | 3 | 334 (2) | 2,122 (3) | 8 |
| Peripheral vascular disease | 889 (6) | 37,031 (7) | 7 | 889 (6) | 6,342 (10) | 17 |
| Rheumatoid arthritis | 218 (1) | 7,181 (1) | 0 | 218 (1) | 1,117 (2) | 3 |
| Renal disease | 1,491 (10) | 51,562 (10) | 2 | 1,491 (10) | 8,706 (14) | 14 |
| Atrial fibrillation | 1,205 (8) | 42,972 (9) | 3 | 1,205 (8) | 6,621 (11) | 10 |
| Arthritis | 1,368 (9) | 48,622 (10) | 3 | 1,368 (9) | 8,294 (14) | 15 |
| Bronchiectasis | 19 (0) | 947 (0) | 2 | 19 (0) | 154 (0) | 3 |
| Depression | 2,254 (15) | 86,456 (17) | 7 | 2,254 (15) | 12,162 (20) | 14 |
| Embolism (history) | 304 (2) | 12,795 (3) | 4 | 304 (2) | 2,164 (4) | 9 |
| Falls (history) | 114 (1) | 6,726 (1) | 6 | 114 (1) | 1,163 (2) | 10 |
| Hepatitis B | 28 (0) | 702 (0) | 1 | 28 (0) | 158 (0) | 2 |
| Hepatitis C | 88 (1) | 3,995 (1) | 3 | 88 (1) | 880 (1) | 9 |
| Hyperlipidemia | 4,628 (30) | 151,284 (30) | 1 | 4,628 (30) | 23,000 (37) | 15 |
| Interstitial lung Disease | 118 (1) | 5,790 (1) | 4 | 118 (1) | 965 (2) | 7 |
| Impaired mobility | 39 (0) | 4,903 (1) | 9 | 39 (0) | 1,026 (2) | 15 |
| Musculoskeletal disorder | 7,390 (48) | 255,372 (51) | 7 | 7,390 (48) | 36,063 (59) | 21 |
| Mycoses | 777 (5) | 37,023 (7) | 10 | 777 (5) | 6,979 (11) | 23 |
| Obesity | 1,710 (11) | 54,124 (11) | 1 | 1,710 (11) | 7,355 (12) | 3 |
| Parkinson's | 117 (1) | 7,521 (1) | 7 | 117 (1) | 1,244 (2) | 11 |
| Sickle cell disease | 11 (0) | 387 (0) | 0 | 11 (0) | 87 (0) | 2 |
| Solid organ transplant recipient | 69 (0) | 2,756 (1) | 2 | 69 (0) | 422 (1) | 3 |
| Stroke / transient ischemic attack | 339 (2) | 16,671 (3) | 7 | 339 (2) | 3,041 (5) | 15 |
| Urinary tract infection | 371 (2) | 19,079 (4) | 8 | 371 (2) | 3,594 (6) | 17 |
| Immunocompromised | 2,324 (15) | 96,783 (20) | 12 | 2,324 (15) | 14,719 (24) | 22 |
| CAN Score, Mean (SD) |  |  |  |  |  |  |
| Mortality-1 year | 5.2% (10) | 5.7% (11) | 5 | 5.2% (10) | 7.6% (12%) | 22 |
| Mortality-90 days | 1.5% (4) | 1.7% (4) | 4 | 1.5% (4) | 2.3% (5%) | 17 |
| Event-1 year | 20.5% (20) | 20.6% (21) | 1 | 20.5% (20) | 24.1% (23%) | 16 |
| Event-90 days | 8.0% (11) | 8.3% (12) | 3 | 8.0% (11) | 10.1% (14%) | 17 |
| Hospitalization-1 year | 17.8% (18) | 18.6% (19) | 4 | 17.8% (18) | 21.6% (21%) | 19 |
| Hospitalization-90 days | 7.2% (10) | 7.8% (11) | 6 | 7.2% (10) | 9.5% (13%) | 20 |
| Any COVID-19 symptoms | 10,499 (68) | 105,464 (21) | 107 | 10,499 (68) | 13,067 (21) | 107 |
| Influenza vaccination (2019/20 season) | 135 (1) | 4,521 (1) | 0 | 135 (1) | 753 (1) | 3 |
| Influenza vaccination (2020/21 season) | 1,570 (10) | 54,432 (11) | 2 | 1,570 (10) | 8,506 (14) | 11 |
| Abbreviations: BMI, body mass index; CAN Score, Care Assessment Needs score; No., number; SMD, standardized mean difference; VA, Veterans Affairs.<br><sup>a</sup>Definitions for variables are provided in Supplemental Table 1.<br><sup>b</sup>SMD of 10 or greater was used to identify imbalance between cases and controls. |  |  |  |  |  |  |

**Figure 1.**
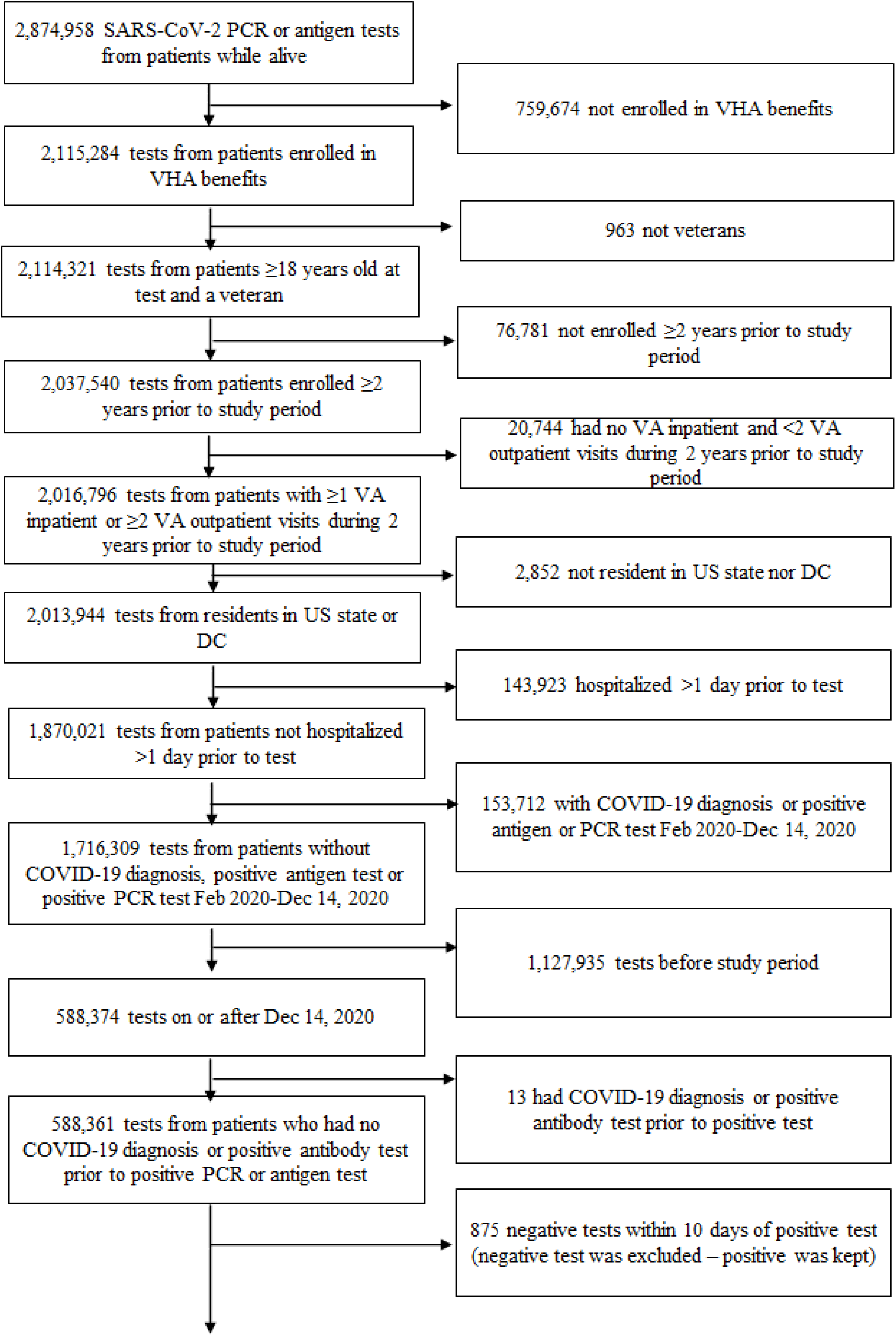
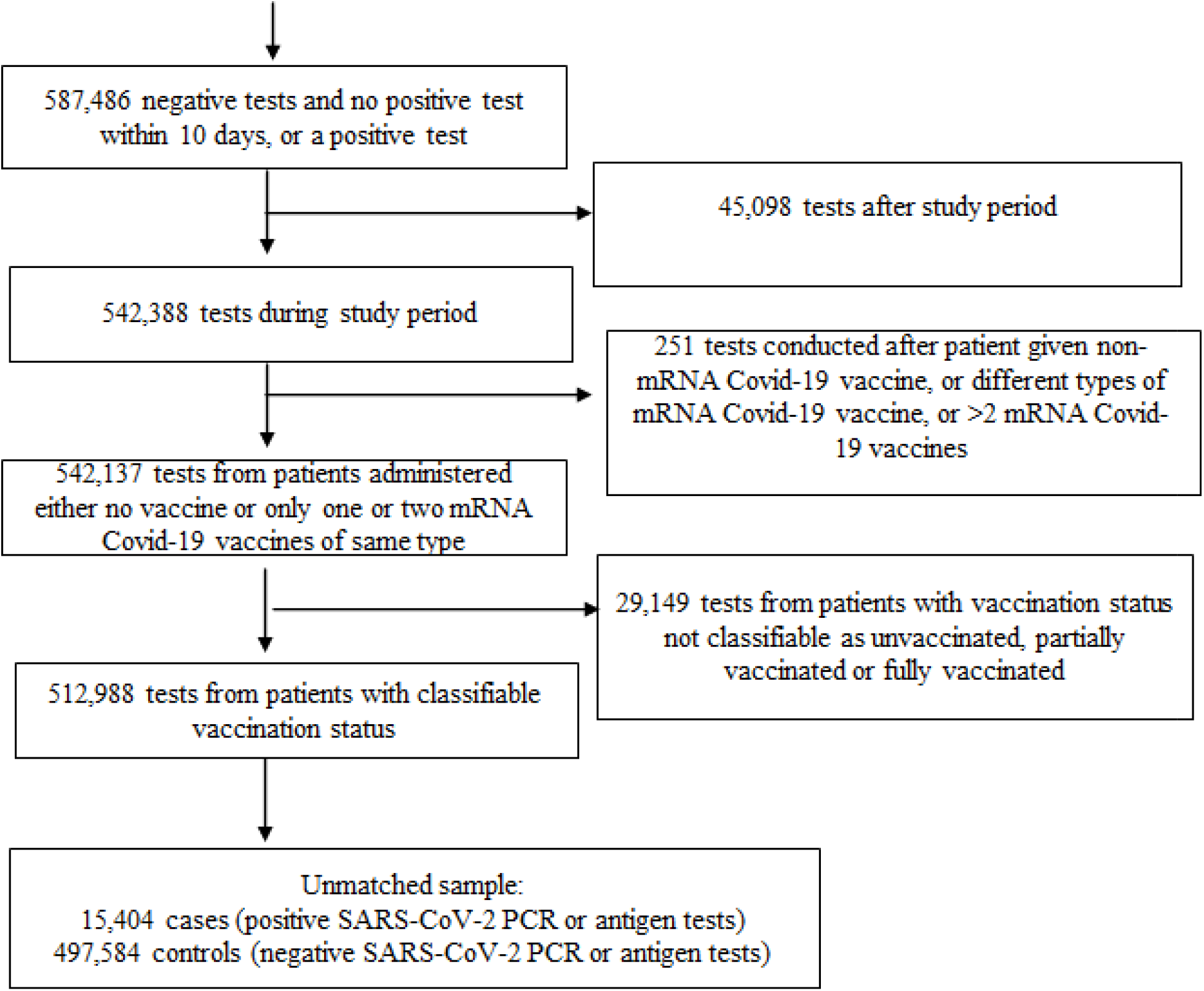
SARS-CoV-2 PCR and antigen tests (December 14, 2020-March 7, 2021) meeting study criteria

### Vaccine Effectiveness

Table 3 shows the estimated VE for full and partial vaccination against a documented positive SARS-CoV-2 test, regardless of symptoms. Among the overall population, the adjusted VE for full and partial vaccinations were 94% (95%CI 92-95) and 58% (95%CI 54-62), respectively. VE was similar to that in the overall population for most subpopulations. VE for full vaccination was a few percentage points lower among patients who were overweight/obese (87% (95%CI 82-91)), had cancer (84% (95%CI 73-91) or had congestive heart disease (85% (95%CI 76-90)).

**Table 3.** Vaccine Effectiveness Against Laboratory Confirmed SARS-CoV-2 Infection^a^

|  | VE, % (95%CI) |  |  |  |
| --- | --- | --- | --- | --- |
|  | Full vs. no vaccination |  | Partial vs. no vaccination |  |
|  | Unadjusted | Adjusted<sup>b</sup> | Unadjusted | Adjusted<sup>b</sup> |
| Overall | 96 (95, 97) | 94 (92, 95) | 70 (67%, 73%) | 58 (54, 62) |
| Age, y |  |  |  |  |
| 18-64 | 91 (84, 95) | 91 (83, 95) | 59 (49, 67) | 52 (40, 62) |
| ≥65 | 92 (90, 94) | 92 (89, 94) | 57 (52, 61) | 54 (48, 58) |
| 18-79 | 91 (88, 93) | 91 (88, 93) | 55 (50, 59) | 53 (48, 58) |
| ≥80 | 93 (89, 96) | 93 (88, 96) | 59 (50, 66) | 56 (46, 64) |
| VHA defined priority group |  |  |  |  |
| 1-4 | 93 (90, 95) | 93 (89, 95) | 60 (53, 65) | 56 (49, 63) |
| 5-6 | 88 (83, 92) | 90 (85, 93) | 53 (45, 60) | 55 (46, 62) |
| 7-8 | 90 (84, 94) | 90 (84, 94) | 43 (33, 52) | 47 (37, 56) |
| Race or ethnic group |  |  |  |  |
| Black | 94 (89, 97) | 93 (88, 96) | 49 (39, 58) | 48 (37, 57) |
| Hispanic | 82 (59, 92) | 73 (38, 89) | 52 (25, 69) | 44 (13, 65) |
| White | 91 (88, 93) | 90 (87, 93) | 56 (51, 61) | 55 (50, 60) |
| Other | 98 (86, 100) | 98 (86, 100) | 61 (40, 75) | 65 (46, 78) |
| Sex |  |  |  |  |
| Female | 86 (62, 95) | 82 (51, 94) | 73 (50, 85) | 65 (35, 81) |
| Male | 92 (90, 94) | 92 (90, 94) | 54 (49, 58) | 54 (50, 58) |
| Urban-rural |  |  |  |  |
| Rural | 93 (89, 96) | 93 (89, 96) | 60 (52, 66) | 58 (50, 65) |
| Urban | 91 (88, 93) | 90 (87, 93) | 52 (47, 57) | 50 (44, 55) |
| Income |  |  |  |  |
| Low income | 92 (83, 96) | 92 (84, 96) | 58 (45, 68) | 56 (42, 66) |
| Homeless |  |  |  |  |
| Homeless | 93 (78, 98) | 93 (79, 98) | 60 (36, 76) | 56 (28, 73) |
| BMI |  |  |  |  |
| Overweight/Obese | 88 (83, 92) | 87 (82, 91) | 48 (41, 55) | 45 (37, 52) |
| Underlying medical conditions |  |  |  |  |
| Cancer | 85 (74, 91) | 84 (73, 91) | 45 (28, 58) | 43 (25, 56) |
| Congestive heart failure | 87 (80, 92) | 85 (76, 90) | 57 (46, 66) | 54 (42, 64) |
| Chronic kidney disease | 93 (87, 96) | 91 (85, 94) | 54 (43, 63) | 56 (45, 64) |
| Diabetes mellitus | 93 (90, 95) | 92 (88, 94) | 59 (53, 65) | 56 (49, 62) |
| Hypertension | 91 (88, 93) | 91 (88, 93) | 56 (50, 61) | 56 (51, 61) |
| Immunocompromised | 86 (79, 90) | 88 (82, 92) | 42 (31, 52) | 43 (32, 52) |
| Adjusted VE, % (95%CI) [reference group: unvaccinated] |  |  |  |  |
|  | Late full vaccination (from 14 days after dose 2) | Full vaccination (from 7 days after dose 2) | 14 to 20 days after dose 1 | Partial vaccination (7 days after dose 1 until dose 2) |
| Overall |  |  |  |  |
|  | 95 (3, 96) | 94 (92, 95) | 63 (57, 69) | 58 (54, 62) |
| BMI |  |  |  |  |
| Overweight/Obese | 91 (85, 95) | 87 (82, 91) | 58 (46, 67) | 45 (37, 52) |
| Underlying medical conditions |  |  |  |  |
| Cancer | 86 (73, 93) | 84 (73, 91) | 56 (29, 73) | 43 (25, 56) |
| Congestive heart failure | 88 (79, 94) | 85 (76, 90) | 70 (53, 81) | 54 (42, 64) |
| Chronic kidney disease | 92 (84 , 95) | 91 (85, 94) | 66 (49, 78) | 55 (45, 64) |
| Diabetes mellitus | 93 (88, 95) | 92 (88, 94) | 64 (53, 72) | 56 (49, 62) |
| Hypertension | 92 (89, 95) | 91 (88, 93) | 62 (53, 69) | 56 (51, 61) |
| Immunocompromised | 86 (78, 91) | 88 (82, 92) | 57 (41, 68) | 43 (32, 52) |

Abbreviations: BMI, body mass index; VE, vaccine effectiveness; VHA, Veterans Health Affairs.
<sup>a</sup>See Supplemental Table 1 for definitions of variables in this table.
<sup>b</sup>The adjusted variables include the following: age, BMI, cancer, congestive heart failure, chronic kidney disease, diabetes mellitus, hypertension, immunocompromised, VA priority level, race, sex, rurality.

VE estimates were similar across all age, sex, race, or urban/rural status, with overlapping 95% CIs. For the overall population and all subpopulations, the VE point estimates for partial vaccination defined by occurrence of cases within 14-20 days after the first dose were higher than those for partial vaccination measured starting 7 days after the first dose. When we restricted the analysis to symptomatic cases, VE for the overall population was 91% (95%CI 87-93), about the same as for all tested individuals (Supplemental Table 2).

Effectiveness estimates against hospitalization (89% (95%CI 81-93)), and against death (98.5% (95%CI 86.6-99.8), were similar to the VE we found against infection and symptomatic disease (Tables 3, 4, Supplemental Table 3).

**Table 4.**
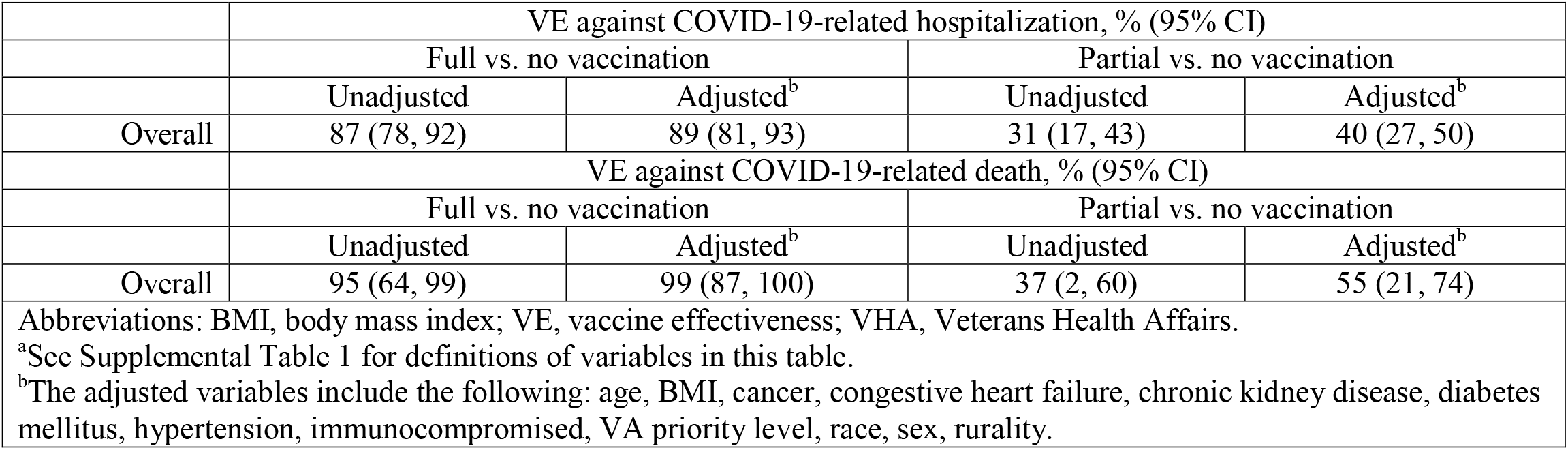
Vaccine Effectiveness Against COVID-19-related Hospitalization and Death^a^

## DISCUSSION

The VHA’s vaccination effort reached all demographic groups among Veterans in the first three months following availability of mRNA COVID vaccines under EUA. Notably, vaccination coverage was at least as high among Black Veterans as White Veterans.

This nationwide study on COVID-19 mRNA vaccines in the US demonstrated high VE of mRNA vaccines against both laboratory confirmed SARS-CoV-2 infections (regardless of symptoms) and symptomatic disease.[Table 3, *Supplement* Table 2] Although our sample size constrained our ability to analyze subpopulations in the short period we investigated, our VE estimates were similarly high in most subpopulations, for symptomatic cases, and against COVID-19 associated hospitalization and death. Our VE estimates are comparable to those from the clinical trials (5, 6) and from other observational studies (Table 3, *Supplement* Table 2). Two VE studies from Israel following the rollout of Pfizer-BioNTech COVID-19 Vaccine among individuals ≥16 years old found similar results: a national surveillance data study, 95.3% [95%CI 94.9-95.7%](7), and a study in Israel’s largest healthcare organization, 92% [95%CI 88-95%](8). Our results were also similar to those from a prospective study among vaccinated healthcare workers in the United Kingdom, 85% [95%CI 74-96%](9), and a smaller study among essential and frontline workers in the US, 90% [95%CI 68-97%](10).

Although power was limited, our VE estimates against COVID-19 hospitalization were comparable to those from two studies in Israel, 97.2% (95%CI 96.8-97.5%) (7) and 87% (95%CI 55-100%)(8), and to a US study of patients ≥65 years old, 94% (95%CI 49-99%)(11) (Table 4). Also comparable were our results for VE against death, which was 96.7% (95%CI 96.0-97.3%) in a study in Israel(7) (Table 4).

Our study, using data from a large and diverse U.S. population, adds important context to our understanding of VE because vaccination rollout and SARS-CoV-2 variants have differed from state to state, and country to country. In Israel and the United Kingdom, mRNA COVID-19 vaccine distribution included only the Pfizer-BioNTech COVID-19 Vaccine, and the United Kingdom extended the interval between doses to vaccinate their population more rapidly with one dose, limiting our ability to directly compare with their results(8, 12). Furthermore, each country has distinct demographic characteristics, so our study allowed for a more in-depth analysis of VE in subpopulations for whom the COVID-19 disease burden has been greater in the U.S., such as Black and Hispanic patients.

Many important prior clinical trials and observational studies (6, 7, 9, 10, 13) had limited sample size to adequately assess the effectiveness of COVID-19 vaccines for people with underlying medical conditions, even though some conditions may predispose individuals to severe consequences from infection(14). The immune response to vaccination among immunocompromised individuals has not been fully explored. We found that, when fully vaccinated, effectiveness is 88% (95%CI, 82-92) for immunocompromised patients, which is reassuring. Further evaluation of these patients by underlying disease and treatment is warranted given the response to infection may vary accordingly (14). Lee et al.(15) demonstrated COVID-19 outcomes may be more severe among patients with hematological malignancies versus solid tumors. As multiple studies, including ours, have shown (5, 9, 10, 12, 13), the mRNA vaccines show some effectiveness even after just the first dose (Tables 3, 4, Supplement Table 2).

COVID-19 has disproportionally affected racial and ethnic minorities and low-income communities (16-18). Our results show the VHA was able to vaccinate minority and low-income Veterans with similar efficiency. The rates of SARS-CoV-2 testing in the first half of 2020 were higher among racial and ethnic minorities than among Whites. Prior to vaccination, African American and Hispanic Veterans were at 30 to 40% higher risk of infection compared to their Caucasian counterparts(17). After VHA’s thorough effort to provide vaccination to all Veterans, regardless of racial/ethnic group or socio-economic status, we found that the risk of SARS-CoV-2 infection (irrespective of symptoms) following full vaccination was reduced for all, with similar VE for all racial and ethnic groups, demonstrating that the equitable distribution of vaccination is an effective means to reduce racial and socio-economic disparity in COVID-19 disease burden.

## STRENGTHS AND LIMITATIONS

A main concern regarding test-negative studies is misclassification. Our study relied on records of vaccination collected prospectively and in near real-time rather than subjects’ recall. While this study demonstrated that vaccination coverage reached all demographic subgroups, the coverage rates reported here may be conservative estimates (19), given that vaccination of individuals by State and local health departments might have not been reported to the VHA. Moreover, by the end of 2020, VHA had standardized its testing and case definitions, unlike at the beginning of the pandemic(20). We used SARS-CoV-2–positive test results coupled with COVID-19 symptoms in a sensitivity analysis to reduce potential misclassification and found that VE matched that of the main analysis at 91% (95%CI 87, 93) (*Supplemental* Table 3). Some Veterans vaccinated through the VHA could have been hospitalized in a non-VHA facility, especially in rural communities (21). We limited the study population to those who routinely sought care at VHA facilities to minimize the likelihood of including patients who were vaccinated or sought treatment for COVID-19 elsewhere. To further address this, we utilized available Medicare data and repeated the analysis including any records of vaccination and hospitalization outside the VHA for these patients. The results were similar with a VE of 87% (95% CI 83, 89) (*Supplemental* Table 4). We also considered that our analysis utilized data from both antigen and PCR tests, and there may be differences in the sensitivities and specificities between tests which could lead to misclassification of cases and controls. We determined that 94% of tests were PCR and 6% were antigen tests. While this misclassification would likely be non-differential, and we repeated the analysis of VE against infection, stratifying by type of test and found no effect on the estimate (*Supplemental* Table 6). Our analysis included test results from all adult patients, without imposing strict rules to identify vaccine eligibility at the time of their test because the prevalence of conditions which would have made patients vaccine eligible early on was quite high (e.g., 55% hypertension among controls); we also conducted analyses for subgroups of patients who would have been vaccine eligible early on. Nevertheless, there could still be residual misclassification and differences in health seeking behavior and disease risk between different subpopulations and for individuals in the VHA and CMS system that would require further investigation in future studies.

## CONCLUSION

Over a period of only three months after the first COVID-19 vaccine was authorized, the VHA successfully vaccinated and tested millions of Veterans of all socio-economic groups for COVID-19. We found mRNA vaccines to be highly effective against the risk of SARS-CoV-2 laboratory-confirmed infection, symptomatic disease, hospitalization, and death. The effectiveness of these vaccines, combined with their equitable and efficient deployment, have resulted in attenuated COVID-19 disease burden among all VHA enrolled Veterans and specific vulnerable populations.

## Data Availability

Because data contain potentially identifying or sensitive patient information, all relevant data must be requested through the Department of Veterans Affairs at:
Research and Development Committee
VA Medical Center
163 Veterans Drive
White River Junction, VT 05009-0001

## Acknowledgements of research support for the study

This project was funded by the United States Food and Drug Administration through an interagency agreement with the Veterans Health Administration

## Supplementary Material

**Supplementary Figure 1.**
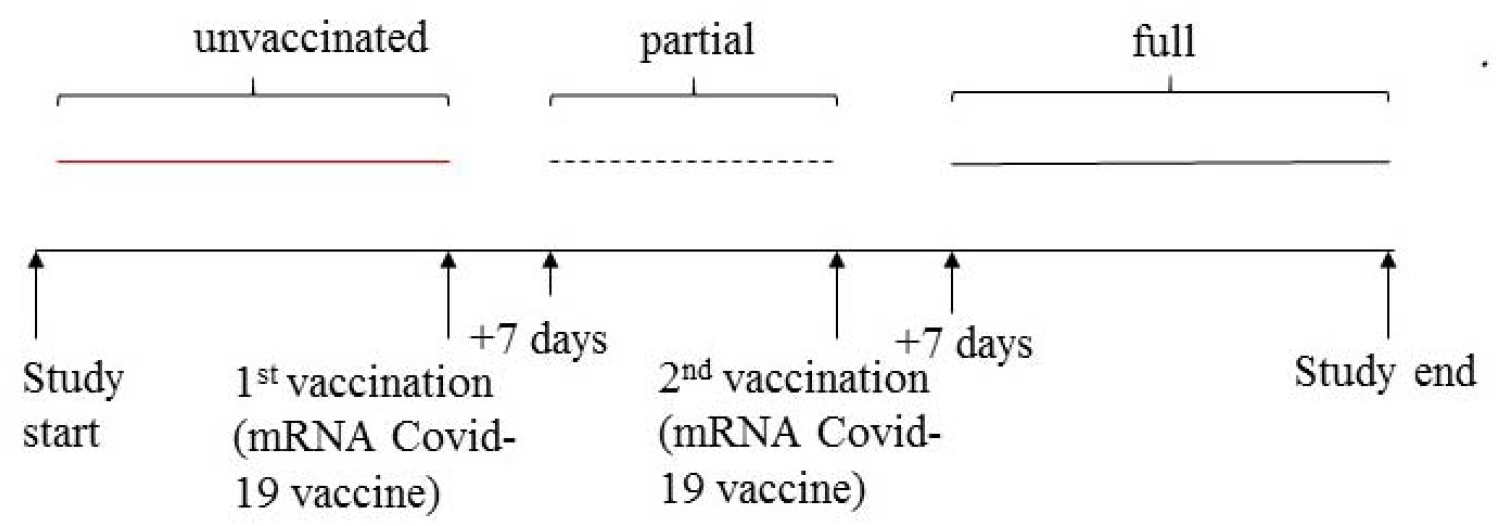
Vaccination exposure classification

**Supplemental Table 1.**
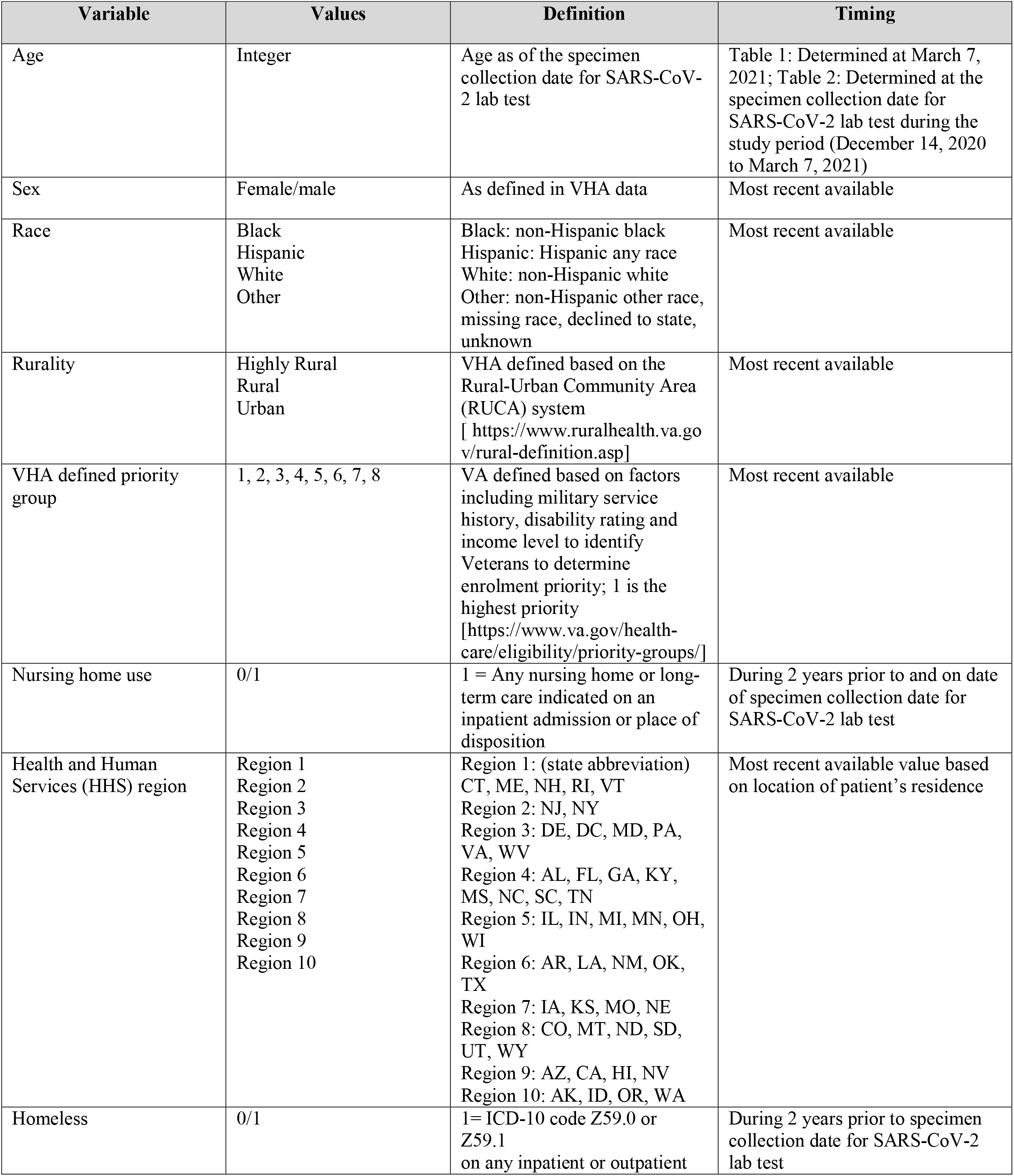
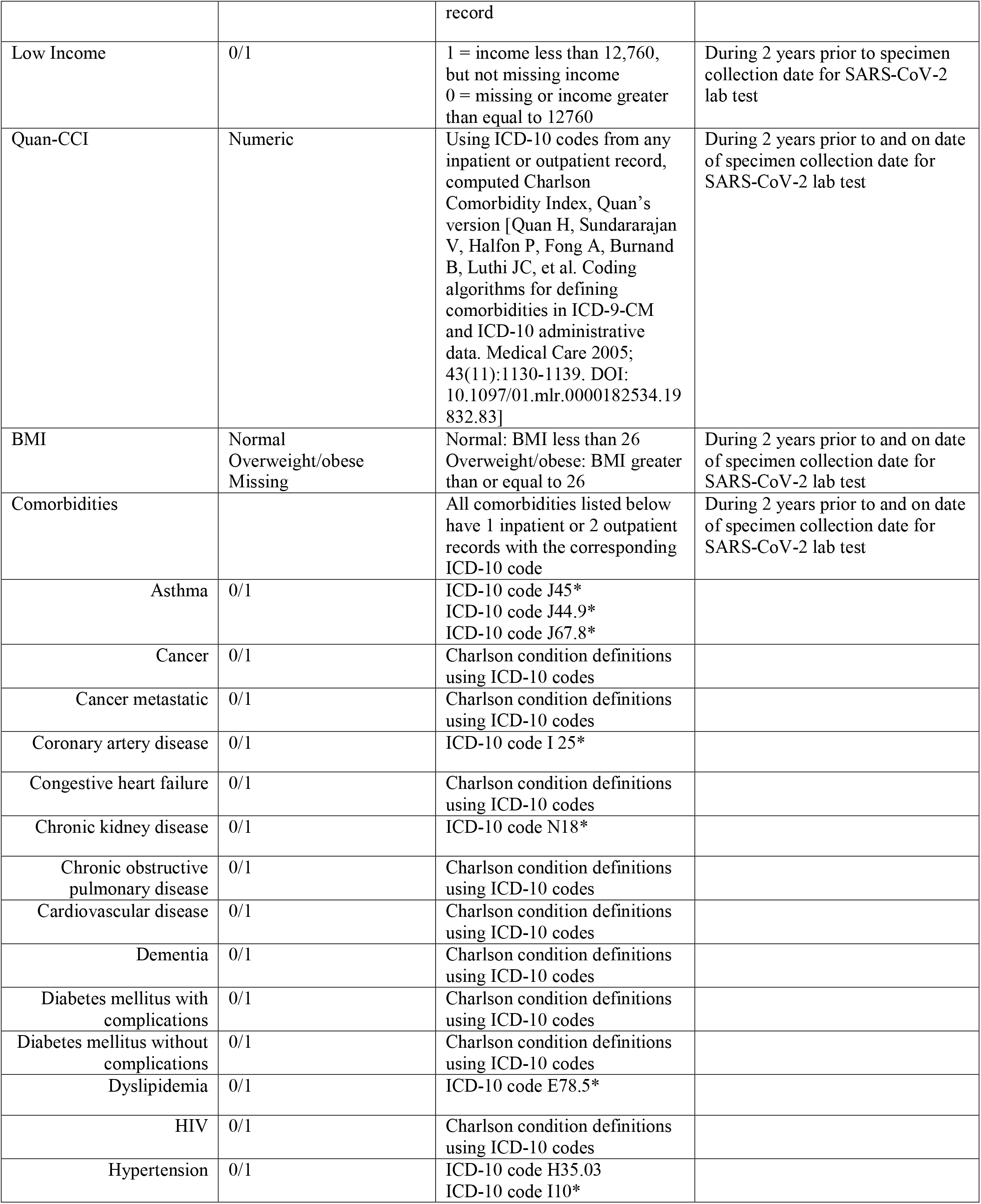
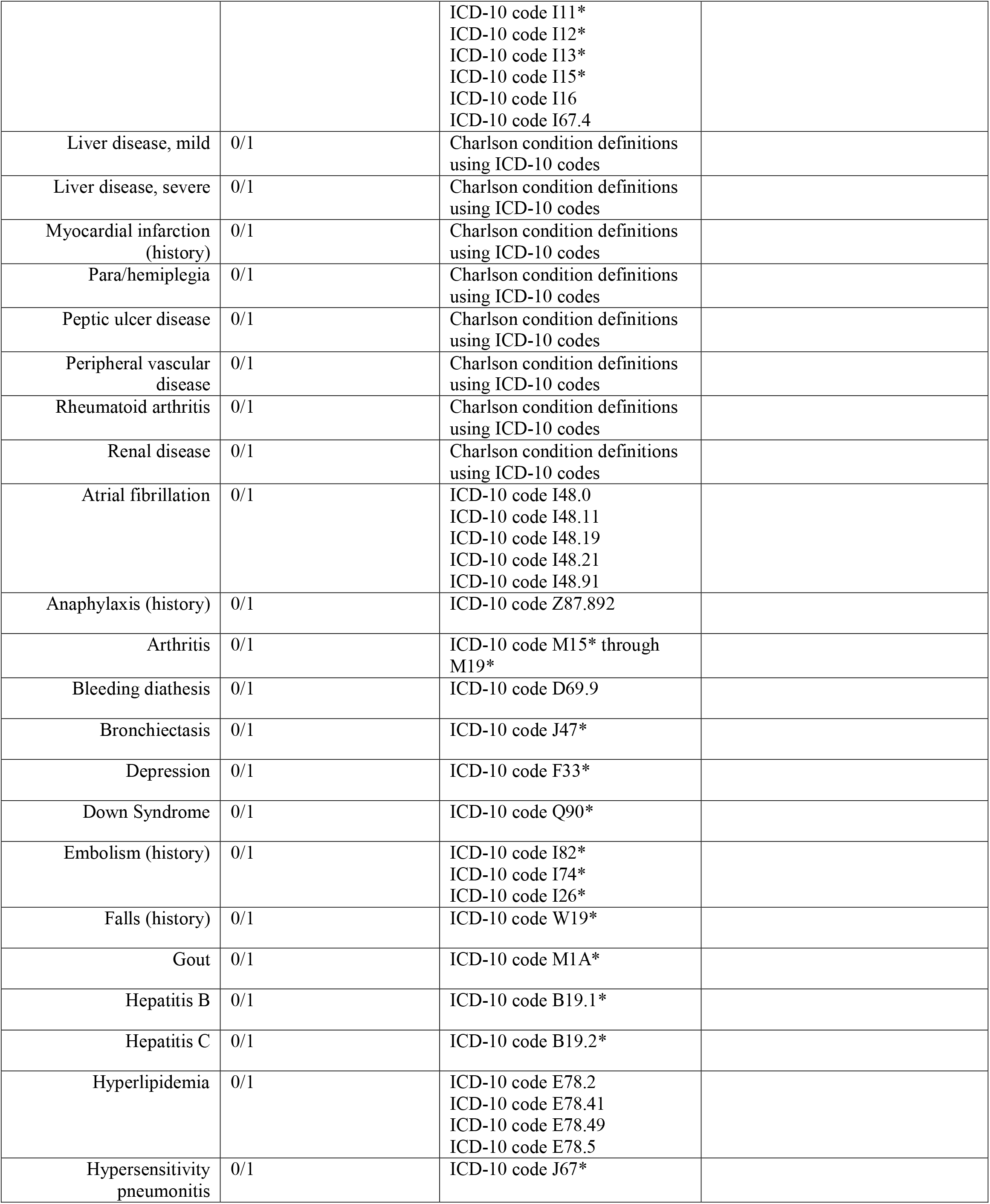
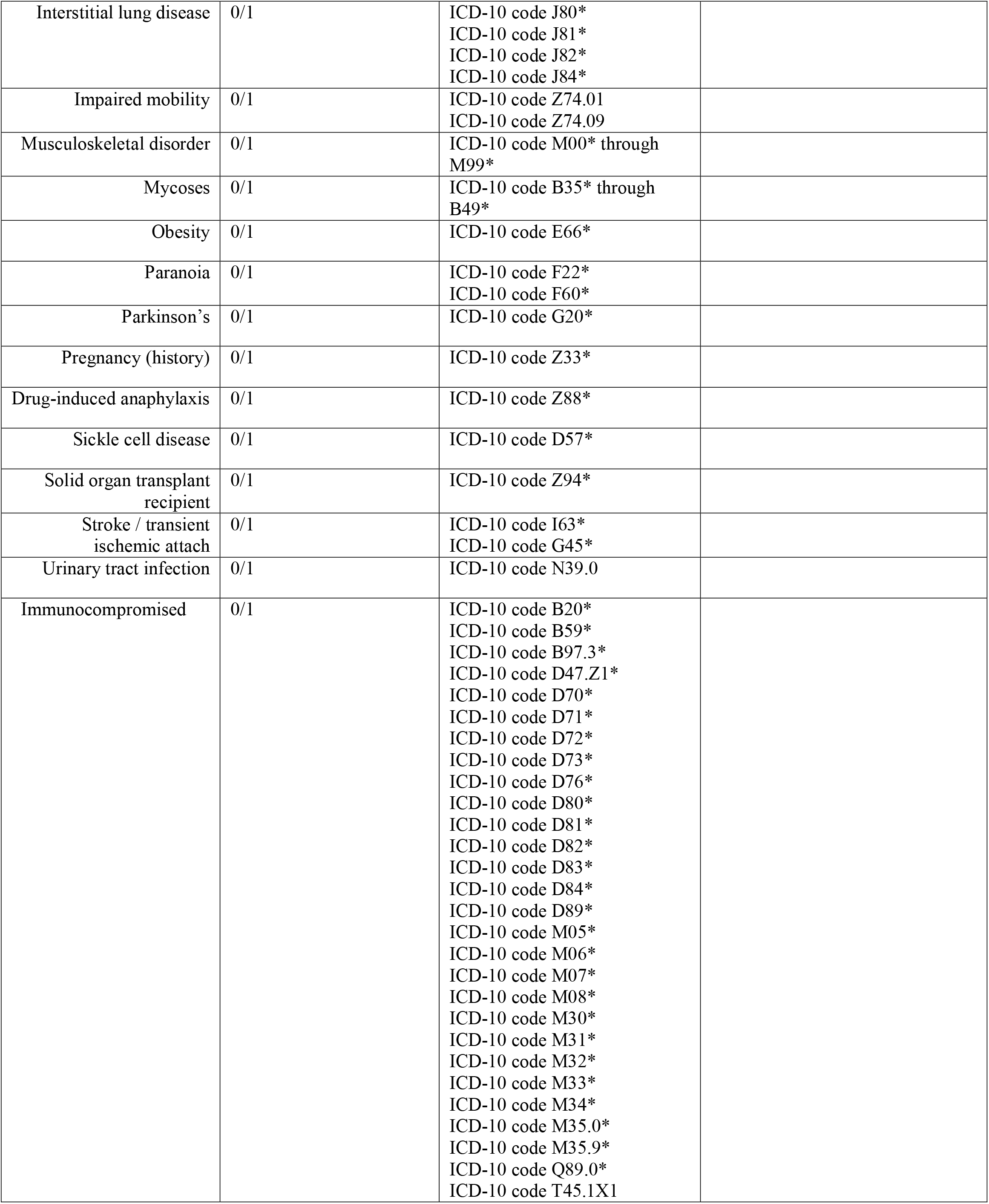
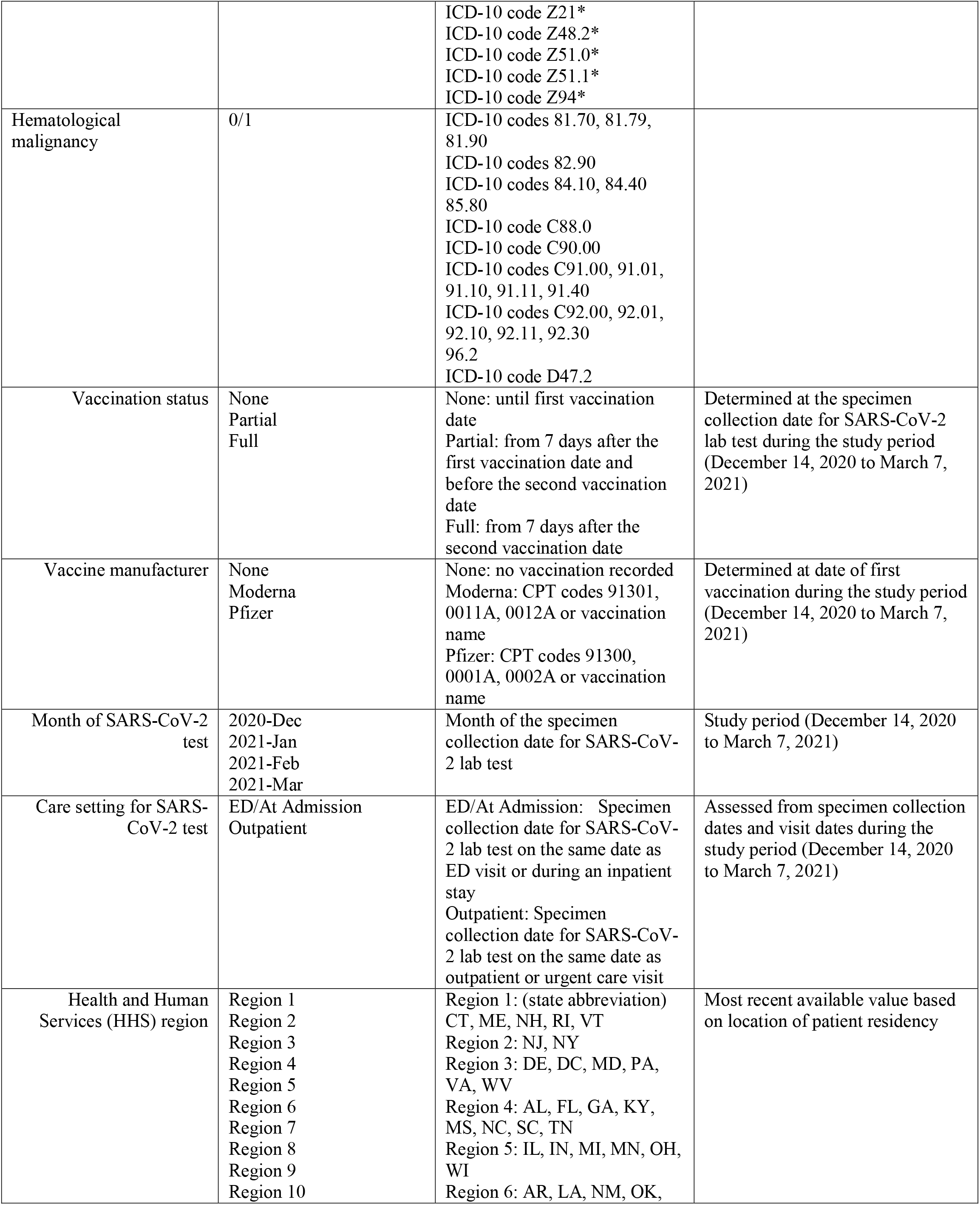
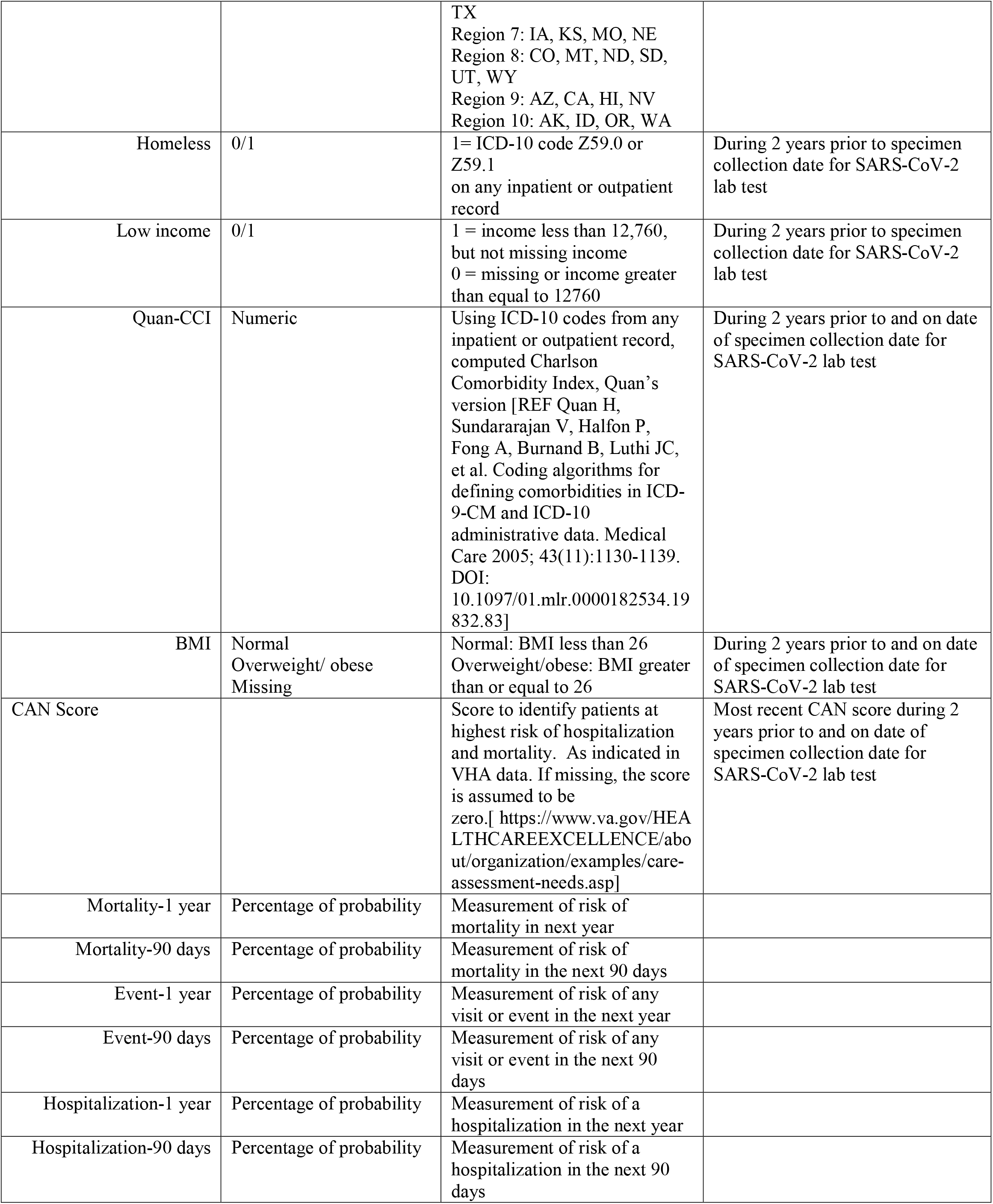
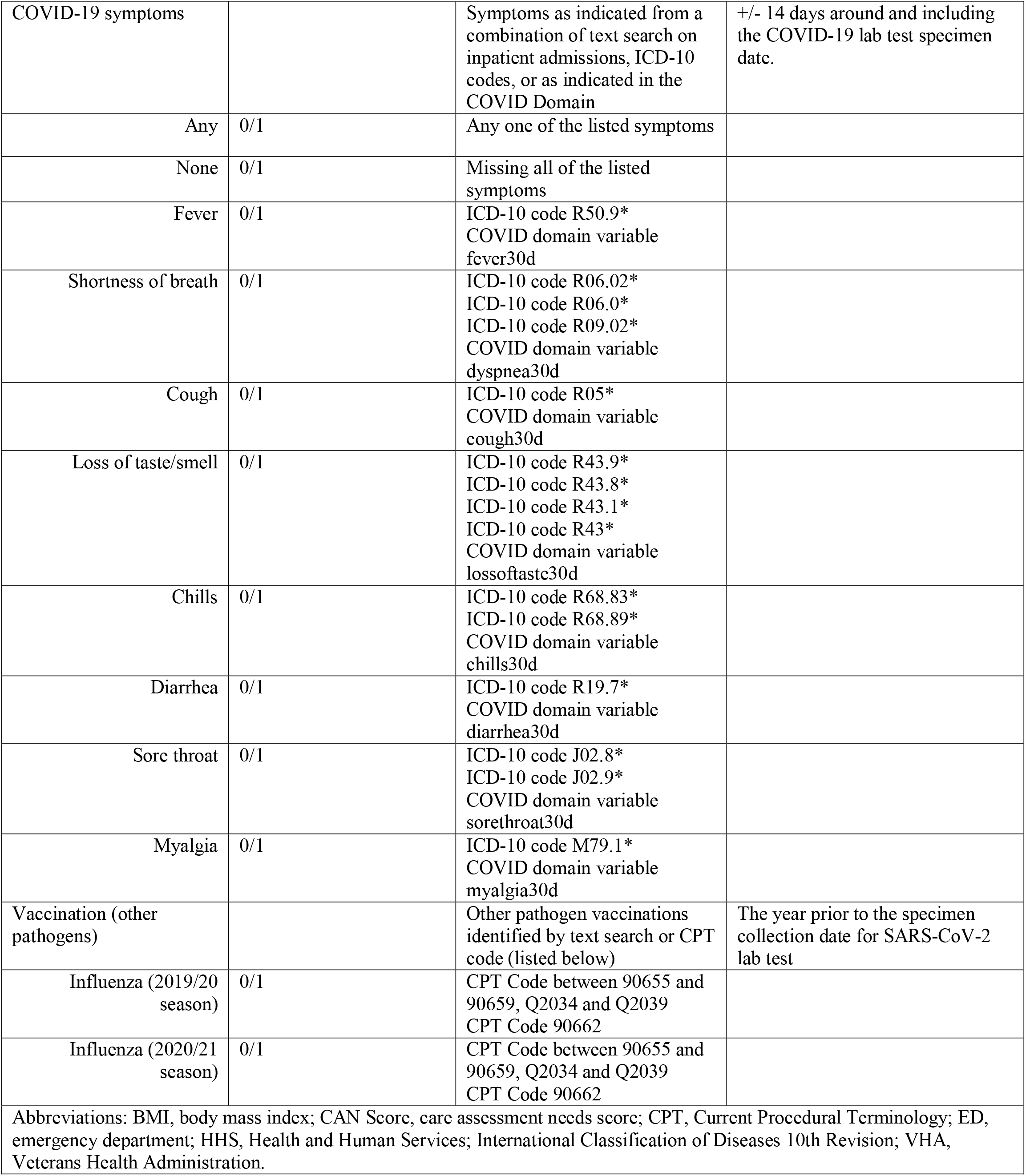
Definitions of variables

| Variable | Values | Definition | Timing |
| --- | --- | --- | --- |
| Age | Integer | Age as of the specimen collection date for SARS-CoV-2 lab test | Table 1: Determined at March 7, 2021; Table 2: Determined at the specimen collection date for SARS-CoV-2 lab test during the study period (December 14, 2020 to March 7, 2021) |
| Sex | Female/male | As defined in VHA data | Most recent available |
| Race | Black<br>Hispanic<br>White<br>Other | Black: non-Hispanic black<br>Hispanic: Hispanic any race<br>White: non-Hispanic white<br>Other: non-Hispanic other race, missing race, declined to state, unknown | Most recent available |
| Rurality | Highly Rural<br>Rural<br>Urban | VHA defined based on the Rural-Urban Community Area (RUCA) system<br>[ https://www.ruralhealth.va.gov/rural-definition.asp] | Most recent available |
| VHA defined priority group | 1, 2, 3, 4, 5, 6, 7, 8 | VA defined based on factors including military service history, disability rating and income level to identify Veterans to determine enrolment priority; 1 is the highest priority<br>[https://www.va.gov/health-care/eligibility/priority-groups/] | Most recent available |
| Nursing home use | 0/1 | 1 = Any nursing home or long-term care indicated on an inpatient admission or place of disposition | During 2 years prior to and on date of specimen collection date for SARS-CoV-2 lab test |
| Health and Human Services (HHS) region | Region 1<br>Region 2<br>Region 3<br>Region 4<br>Region 5<br>Region 6<br>Region 7<br>Region 8<br>Region 9<br>Region 10 | Region 1: (state abbreviation) CT, ME, NH, RI, VT<br>Region 2: NJ, NY<br>Region 3: DE, DC, MD, PA, VA, WV<br>Region 4: AL, FL, GA, KY, MS, NC, SC, TN<br>Region 5: IL, IN, MI, MN, OH, WI<br>Region 6: AR, LA, NM, OK, TX<br>Region 7: IA, KS, MO, NE<br>Region 8: CO, MT, ND, SD, UT, WY<br>Region 9: AZ, CA, HI, NV<br>Region 10: AK, ID, OR, WA | Most recent available value based on location of patient's residence |
| Homeless | 0/1 | 1= ICD-10 code Z59.0 or<br>Z59.1<br>on any inpatient or outpatient<br>record | During 2 years prior to specimen<br>collection date for SARS-CoV-2<br>lab test |
| Low Income | 0/1 | 1 = income less than 12,760,<br>but not missing income<br>0 = missing or income greater<br>than equal to 12760 | During 2 years prior to specimen<br>collection date for SARS-CoV-2<br>lab test |
| Quan-CCI | Numeric | Using ICD-10 codes from any<br>inpatient or outpatient record,<br>computed Charlson<br>Comorbidity Index, Quan's<br>version [Quan H, Sundararajan<br>V, Halfon P, Fong A, Burnand<br>B, Luthi JC, et al. Coding<br>algorithms for defining<br>comorbidities in ICD-9-CM<br>and ICD-10 administrative<br>data. Medical Care 2005;<br>43(11):1130-1139. DOI:<br>10.1097/01.mlr.0000182534.19<br>832.83] | During 2 years prior to and on date<br>of specimen collection date for<br>SARS-CoV-2 lab test |
| BMI | Normal<br>Overweight/ obese<br>Missing | Normal: BMI less than 26<br>Overweight/obese: BMI greater<br>than or equal to 26 | During 2 years prior to and on date<br>of specimen collection date for<br>SARS-CoV-2 lab test |
| Comorbidities |  | All comorbidities listed below<br>have 1 inpatient or 2 outpatient<br>records with the corresponding<br>ICD-10 code | During 2 years prior to and on date<br>of specimen collection date for<br>SARS-CoV-2 lab test |
| Asthma | 0/1 | ICD-10 code J45*<br>ICD-10 code J44.9*<br>ICD-10 code J67.8* |  |
| Cancer | 0/1 | Charlson condition definitions<br>using ICD-10 codes |  |
| Cancer metastatic | 0/1 | Charlson condition definitions<br>using ICD-10 codes |  |
| Coronary artery disease | 0/1 | ICD-10 code I 25* |  |
| Congestive heart failure | 0/1 | Charlson condition definitions<br>using ICD-10 codes |  |
| Chronic kidney disease | 0/1 | ICD-10 code N18* |  |
| Chronic obstructive<br>pulmonary disease | 0/1 | Charlson condition definitions<br>using ICD-10 codes |  |
| Cardiovascular disease | 0/1 | Charlson condition definitions<br>using ICD-10 codes |  |
| Dementia | 0/1 | Charlson condition definitions<br>using ICD-10 codes |  |
| Diabetes mellitus with<br>complications | 0/1 | Charlson condition definitions<br>using ICD-10 codes |  |
| Diabetes mellitus without<br>complications | 0/1 | Charlson condition definitions<br>using ICD-10 codes |  |
| Dyslipidemia | 0/1 | ICD-10 code E78.5* |  |
| HIV | 0/1 | Charlson condition definitions<br>using ICD-10 codes |  |
| Hypertension | 0/1 | ICD-10 code H35.03<br>ICD-10 code I10* |  |
|  |  | ICD-10 code I11*<br>ICD-10 code I12*<br>ICD-10 code I13*<br>ICD-10 code I15*<br>ICD-10 code I16<br>ICD-10 code I67.4 |  |
| Liver disease, mild | 0/1 | Charlson condition definitions using ICD-10 codes |  |
| Liver disease, severe | 0/1 | Charlson condition definitions using ICD-10 codes |  |
| Myocardial infarction (history) | 0/1 | Charlson condition definitions using ICD-10 codes |  |
| Para/hemiplegia | 0/1 | Charlson condition definitions using ICD-10 codes |  |
| Peptic ulcer disease | 0/1 | Charlson condition definitions using ICD-10 codes |  |
| Peripheral vascular disease | 0/1 | Charlson condition definitions using ICD-10 codes |  |
| Rheumatoid arthritis | 0/1 | Charlson condition definitions using ICD-10 codes |  |
| Renal disease | 0/1 | Charlson condition definitions using ICD-10 codes |  |
| Atrial fibrillation | 0/1 | ICD-10 code I48.0<br>ICD-10 code I48.11<br>ICD-10 code I48.19<br>ICD-10 code I48.21<br>ICD-10 code I48.91 |  |
| Anaphylaxis (history) | 0/1 | ICD-10 code Z87.892 |  |
| Arthritis | 0/1 | ICD-10 code M15* through M19* |  |
| Bleeding diathesis | 0/1 | ICD-10 code D69.9 |  |
| Bronchiectasis | 0/1 | ICD-10 code J47* |  |
| Depression | 0/1 | ICD-10 code F33* |  |
| Down Syndrome | 0/1 | ICD-10 code Q90* |  |
| Embolism (history) | 0/1 | ICD-10 code I82*<br>ICD-10 code I74*<br>ICD-10 code I26* |  |
| Falls (history) | 0/1 | ICD-10 code W19* |  |
| Gout | 0/1 | ICD-10 code M1A* |  |
| Hepatitis B | 0/1 | ICD-10 code B19.1* |  |
| Hepatitis C | 0/1 | ICD-10 code B19.2* |  |
| Hyperlipidemia | 0/1 | ICD-10 code E78.2<br>ICD-10 code E78.41<br>ICD-10 code E78.49<br>ICD-10 code E78.5 |  |
| Hypersensitivity pneumonitis | 0/1 | ICD-10 code J67* |  |
| Interstitial lung disease | 0/1 | ICD-10 code J80*<br>ICD-10 code J81*<br>ICD-10 code J82*<br>ICD-10 code J84* |  |
| Impaired mobility | 0/1 | ICD-10 code Z74.01<br>ICD-10 code Z74.09 |  |
| Musculoskeletal disorder | 0/1 | ICD-10 code M00* through<br>M99* |  |
| Mycoses | 0/1 | ICD-10 code B35* through<br>B49* |  |
| Obesity | 0/1 | ICD-10 code E66* |  |
| Paranoia | 0/1 | ICD-10 code F22*<br>ICD-10 code F60* |  |
| Parkinson's | 0/1 | ICD-10 code G20* |  |
| Pregnancy (history) | 0/1 | ICD-10 code Z33* |  |
| Drug-induced anaphylaxis | 0/1 | ICD-10 code Z88* |  |
| Sickle cell disease | 0/1 | ICD-10 code D57* |  |
| Solid organ transplant<br>recipient | 0/1 | ICD-10 code Z94* |  |
| Stroke / transient<br>ischemic attach | 0/1 | ICD-10 code I63*<br>ICD-10 code G45* |  |
| Urinary tract infection | 0/1 | ICD-10 code N39.0 |  |
| Immunocompromised | 0/1 | ICD-10 code B20*<br>ICD-10 code B59*<br>ICD-10 code B97.3*<br>ICD-10 code D47.Z1*<br>ICD-10 code D70*<br>ICD-10 code D71*<br>ICD-10 code D72*<br>ICD-10 code D73*<br>ICD-10 code D76*<br>ICD-10 code D80*<br>ICD-10 code D81*<br>ICD-10 code D82*<br>ICD-10 code D83*<br>ICD-10 code D84*<br>ICD-10 code D89*<br>ICD-10 code M05*<br>ICD-10 code M06*<br>ICD-10 code M07*<br>ICD-10 code M08*<br>ICD-10 code M30*<br>ICD-10 code M31*<br>ICD-10 code M32*<br>ICD-10 code M33*<br>ICD-10 code M34*<br>ICD-10 code M35.0*<br>ICD-10 code M35.9*<br>ICD-10 code Q89.0*<br>ICD-10 code T45.1X1 |  |
|  |  | ICD-10 code Z21*<br>ICD-10 code Z48.2*<br>ICD-10 code Z51.0*<br>ICD-10 code Z51.1*<br>ICD-10 code Z94* |  |
| Hematological malignancy | 0/1 | ICD-10 codes 81.70, 81.79, 81.90<br>ICD-10 codes 82.90<br>ICD-10 codes 84.10, 84.40<br>85.80<br>ICD-10 code C88.0<br>ICD-10 code C90.00<br>ICD-10 codes C91.00, 91.01, 91.10, 91.11, 91.40<br>ICD-10 codes C92.00, 92.01, 92.10, 92.11, 92.30<br>96.2<br>ICD-10 code D47.2 |  |
| Vaccination status | None<br>Partial<br>Full | None: until first vaccination date<br>Partial: from 7 days after the first vaccination date and before the second vaccination date<br>Full: from 7 days after the second vaccination date | Determined at the specimen collection date for SARS-CoV-2 lab test during the study period (December 14, 2020 to March 7, 2021) |
| Vaccine manufacturer | None<br>Moderna<br>Pfizer | None: no vaccination recorded<br>Moderna: CPT codes 91301, 0011A, 0012A or vaccination name<br>Pfizer: CPT codes 91300, 0001A, 0002A or vaccination name | Determined at date of first vaccination during the study period (December 14, 2020 to March 7, 2021) |
| Month of SARS-CoV-2 test | 2020-Dec<br>2021-Jan<br>2021-Feb<br>2021-Mar | Month of the specimen collection date for SARS-CoV-2 lab test | Study period (December 14, 2020 to March 7, 2021) |
| Care setting for SARS-CoV-2 test | ED/At Admission<br>Outpatient | ED/At Admission: Specimen collection date for SARS-CoV-2 lab test on the same date as ED visit or during an inpatient stay<br>Outpatient: Specimen collection date for SARS-CoV-2 lab test on the same date as outpatient or urgent care visit | Assessed from specimen collection dates and visit dates during the study period (December 14, 2020 to March 7, 2021) |
| Health and Human Services (HHS) region | Region 1<br>Region 2<br>Region 3<br>Region 4<br>Region 5<br>Region 6<br>Region 7<br>Region 8<br>Region 9<br>Region 10 | Region 1: (state abbreviation) CT, ME, NH, RI, VT<br>Region 2: NJ, NY<br>Region 3: DE, DC, MD, PA, VA, WV<br>Region 4: AL, FL, GA, KY, MS, NC, SC, TN<br>Region 5: IL, IN, MI, MN, OH, WI<br>Region 6: AR, LA, NM, OK, | Most recent available value based on location of patient residency |
|  |  | TX<br>Region 7: IA, KS, MO, NE<br>Region 8: CO, MT, ND, SD,<br>UT, WY<br>Region 9: AZ, CA, HI, NV<br>Region 10: AK, ID, OR, WA |  |
| Homeless | 0/1 | 1= ICD-10 code Z59.0 or<br>Z59.1<br>on any inpatient or outpatient<br>record | During 2 years prior to specimen<br>collection date for SARS-CoV-2<br>lab test |
| Low income | 0/1 | 1 = income less than 12,760,<br>but not missing income<br>0 = missing or income greater<br>than equal to 12760 | During 2 years prior to specimen<br>collection date for SARS-CoV-2<br>lab test |
| Quan-CCI | Numeric | Using ICD-10 codes from any<br>inpatient or outpatient record,<br>computed Charlson<br>Comorbidity Index, Quan's<br>version [REF Quan H,<br>Sundararajan V, Halfon P,<br>Fong A, Burnand B, Luthi JC,<br>et al. Coding algorithms for<br>defining comorbidities in ICD-9-CM and ICD-10<br>administrative data. Medical<br>Care 2005; 43(11):1130-1139.<br>DOI:<br>10.1097/01.mlr.0000182534.19<br>832.83] | During 2 years prior to and on date<br>of specimen collection date for<br>SARS-CoV-2 lab test |
| BMI | Normal<br>Overweight/ obese<br>Missing | Normal: BMI less than 26<br>Overweight/obese: BMI greater<br>than or equal to 26 | During 2 years prior to and on date<br>of specimen collection date for<br>SARS-CoV-2 lab test |
| CAN Score |  | Score to identify patients at<br>highest risk of hospitalization<br>and mortality. As indicated in<br>VHA data. If missing, the score<br>is assumed to be<br>zero.[ https://www.va.gov/HEALTHCAREEXCELLENCE/about/organization/examples/care-assessment-needs.asp] | Most recent CAN score during 2<br>years prior to and on date of<br>specimen collection date for<br>SARS-CoV-2 lab test |
| Mortality-1 year | Percentage of probability | Measurement of risk of<br>mortality in next year |  |
| Mortality-90 days | Percentage of probability | Measurement of risk of<br>mortality in the next 90 days |  |
| Event-1 year | Percentage of probability | Measurement of risk of any<br>visit or event in the next year |  |
| Event-90 days | Percentage of probability | Measurement of risk of any<br>visit or event in the next 90<br>days |  |
| Hospitalization-1 year | Percentage of probability | Measurement of risk of a<br>hospitalization in the next year |  |
| Hospitalization-90 days | Percentage of probability | Measurement of risk of a<br>hospitalization in the next 90<br>days |  |
| COVID-19 symptoms |  | Symptoms as indicated from a combination of text search on inpatient admissions, ICD-10 codes, or as indicated in the COVID Domain | +/- 14 days around and including the COVID-19 lab test specimen date. |
| Any | 0/1 | Any one of the listed symptoms |  |
| None | 0/1 | Missing all of the listed symptoms |  |
| Fever | 0/1 | ICD-10 code R50.9*<br>COVID domain variable fever30d |  |
| Shortness of breath | 0/1 | ICD-10 code R06.02*<br>ICD-10 code R06.0*<br>ICD-10 code R09.02*<br>COVID domain variable dyspnea30d |  |
| Cough | 0/1 | ICD-10 code R05*<br>COVID domain variable cough30d |  |
| Loss of taste/smell | 0/1 | ICD-10 code R43.9*<br>ICD-10 code R43.8*<br>ICD-10 code R43.1*<br>ICD-10 code R43*<br>COVID domain variable lossoftaste30d |  |
| Chills | 0/1 | ICD-10 code R68.83*<br>ICD-10 code R68.89*<br>COVID domain variable chills30d |  |
| Diarrhea | 0/1 | ICD-10 code R19.7*<br>COVID domain variable diarrhea30d |  |
| Sore throat | 0/1 | ICD-10 code J02.8*<br>ICD-10 code J02.9*<br>COVID domain variable sorethroat30d |  |
| Myalgia | 0/1 | ICD-10 code M79.1*<br>COVID domain variable myalgia30d |  |
| Vaccination (other pathogens) |  | Other pathogen vaccinations identified by text search or CPT code (listed below) | The year prior to the specimen collection date for SARS-CoV-2 lab test |
| Influenza (2019/20 season) | 0/1 | CPT Code between 90655 and 90659, Q2034 and Q2039<br>CPT Code 90662 |  |
| Influenza (2020/21 season) | 0/1 | CPT Code between 90655 and 90659, Q2034 and Q2039<br>CPT Code 90662 |  |
| Abbreviations: BMI, body mass index; CAN Score, care assessment needs score; CPT, Current Procedural Terminology; ED, emergency department; HHS, Health and Human Services; International Classification of Diseases 10th Revision; VHA, Veterans Health Administration. |  |  |  |

**Supplemental Table 2.**
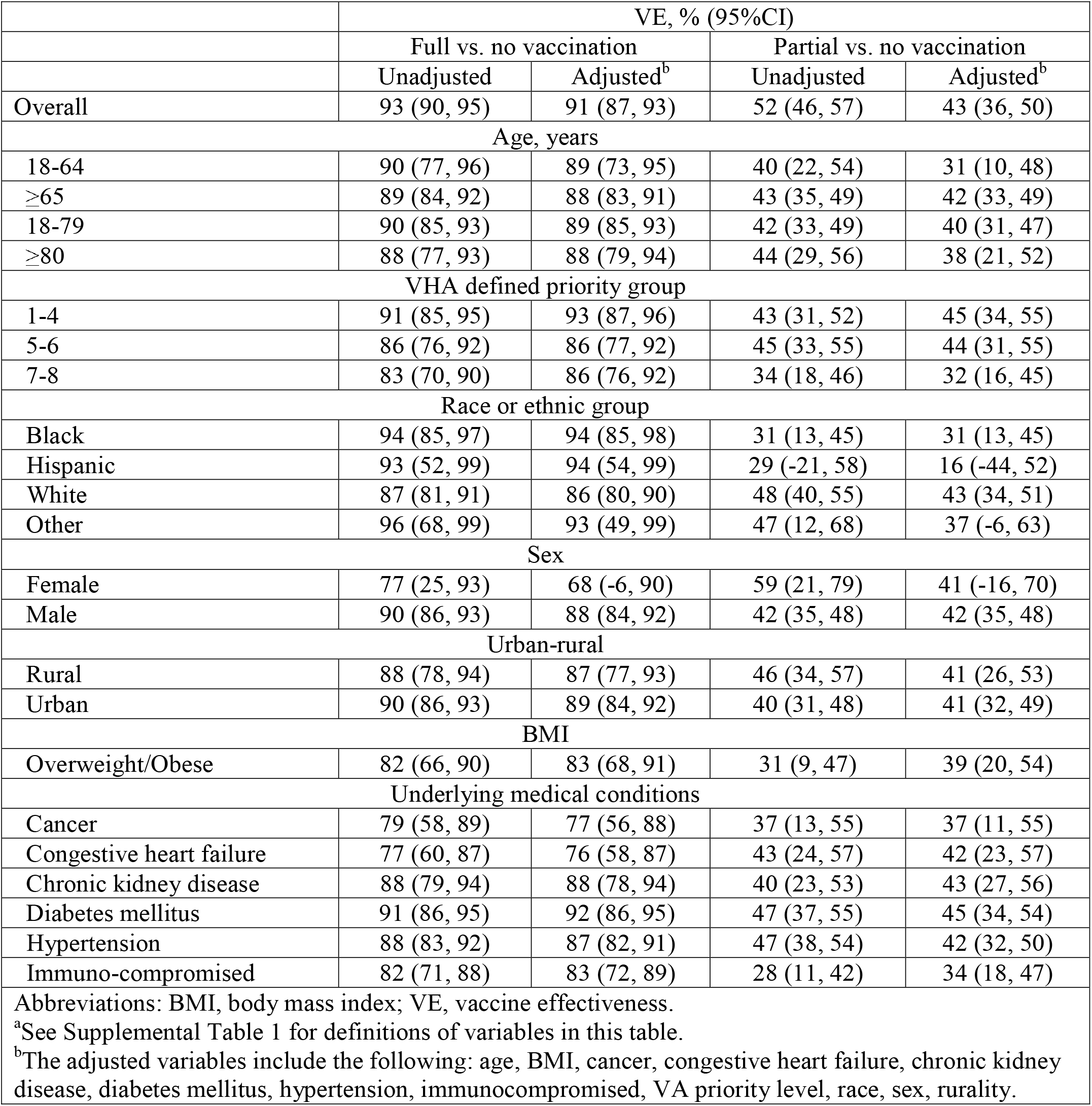
Vaccine Effectiveness Against Laboratory Confirmed SARS-CoV-2 Infection Among Patients with COVID-19 Symptoms^a^

**Supplemental Table 3.** Vaccination Status for Cases and Controls in Analysis of Vaccine Effectiveness Against COVID-19-related Hospitalization and Death

| COVID-19-related hospitalization |  |  |  |  |
| --- | --- | --- | --- | --- |
|  | Unmatched |  | Matched |  |
|  | Cases<br>(n=3,209) | Controls<br>(n=316,895) | Cases<br>(n=3,209) | Controls<br>(n=13,075) |
| COVID-19 vaccination status |  |  |  |  |
| Full | 17 | 11,656 | 17 | 442 |
| Partial | 151 | 18,901 | 151 | 843 |
| None | 3,041 | 286,338 | 3,041 | 11,790 |
| COVID-19-related death |  |  |  |  |
|  | Unmatched |  | Matched |  |
|  | Cases<br>(n=669) | Controls<br>(n=316,895) | Cases<br>(n=669) | Controls<br>(n=2,728) |
| COVID-19 vaccination status |  |  |  |  |
| Vaccinated* | 25 | 30,557 | 25 | 215 |
| None | 644 | 286,338 | 644 | 2,513 |
| *A breakdown by full and partial vaccination status would violate federal cell size suppression policy |  |  |  |  |

**Supplemental Table 4.** Vaccine Effectiveness Against Laboratory Confirmed SARS-CoV-2 Infection, Combining VA and CMS Data^a,b^

|  | VE, % (95%CI) |  |  |  |
| --- | --- | --- | --- | --- |
|  | Full vs. no vaccination |  | Partial vs. no vaccination |  |
|  | Unadjusted | Adjusted<sup>c</sup> | Unadjusted | Adjusted<sup>c</sup> |
| Overall | 87 (83, 89) | 87 (83, 89) | 43 (37, 48) | 42 (36, 48) |
| Age, y |  |  |  |  |
| 18-64 | 89 (77, 95) | 90 (79, 95) | 44 (27, 57) | 43 (26, 57) |
| ≥65 | 88 (84, 91) | 88 (84, 91) | 47 (41, 53) | 46 (39, 51) |
| VHA defined priority group |  |  |  |  |
| 1-4 | 87 (79, 92) | 87 (80, 92) | 52 (42, 61) | 51 (40, 60) |
| 5-6 | 84 (72, 90) | 83 (71, 90) | 44 (30, 56) | 45 (30, 56) |
| 7-8 | 85 (69, 93) | 83 (64, 92) | 22 (-3, 41) | 24 (-2, 43) |
| Race or ethnic group |  |  |  |  |
| Black | 92 (82, 96) | 91 (81, 96) | 37 (16, 52) | 30 (7, 47) |
| White | 83 (78, 88) | 84 (78, 88) | 44 (36, 51) | 46 (39, 53) |
| Sex |  |  |  |  |
| Male | 87 (83, 90) | 87 (83, 90) | 42 (36, 48) | 41 (34, 46) |
| Urban-rural |  |  |  |  |
| Rural | 87 (76, 93) | 87 (76, 93) | 45 (30, 57) | 46 (31, 58) |
| Urban | 84 (78, 88) | 83 (78, 88) | 38 (30, 45) | 36 (27, 44) |
| BMI |  |  |  |  |
| Obese | 84 (75, 90) | 83 (74, 89) | 29 (15, 40) | 27 (12, 39) |
| Underlying medical conditions |  |  |  |  |
| Congestive heart failure | 67 (10, 88) | 71 (17, 90) | 47 (8, 70) | 49 (9, 72) |
| Chronic kidney disease | 86 (53, 96) | 87 (56, 96) | 51 (23, 69) | 47 (15, 67) |
| Diabetes mellitus | 90 (81, 94) | 89 (80, 94) | 36 (20, 49) | 34 (17, 48) |
| Hypertension | 85 (78, 89) | 85 (79, 90) | 35 (24, 45) | 36 (24, 45) |
| Immunocompromised | 65 (32, 82) | 67 (35, 83) | 33 (6, 52) | 37 (11, 55) |

Abbreviations: BMI, body mass index; CMS, Centers for Medicare and Medicaid Services; ICD-10, International Classification of Diseases 10<sup>th</sup> Revision; VA, Veterans Affairs; VE, vaccine effectiveness.
<sup>a</sup>Since 2020, for VHA enrolled patients with Medicare who have either tested positive for SARS-Cov-2 at a VHA facility or who have a COVID-19 medical claim in Medicare (ICD-10 codes B97.29 [“Other coronavirus”] and U07.1 [“COVID-19”] diagnosis code in inpatient, skilled nursing facility, institutional outpatient, hospice, and carrier/Part B files), the VHA receives the patients’ Medicare data with a 4-5 week lag time [REF https://vaww.virec.research.va.gov/VACMS/Medicare/COVID19-Data.htm].
<sup>b</sup>See Supplemental Table 1 for definitions of variables in this table.
<sup>c</sup>The adjusted variables include the following: age, BMI, cancer, congestive heart failure, chronic kidney disease, diabetes mellitus, hypertension, immunocompromised, VA priority level, race, sex, rurality.

**Supplemental Table 5.** Vaccine Effectiveness Against Laboratory Confirmed SARS-CoV-2 Infection by Type of Diagnostic Test^a^

|  | VE, % (95% CI) |  |  |  |
| --- | --- | --- | --- | --- |
| Overall | Full vs. no vaccination |  | Partial vs. no vaccination |  |
|  | Unadjusted | Adjusted<sup>b</sup> | Unadjusted | Adjusted<sup>b</sup> |
| PCR test | 95 (93, 96) | 92 (89, 94) | 68 (64, 70) | 53 (49, 58) |
| Antigen test | 96 (90, 99) | 97 (90, 99) | 54 (33, 69) | 62 (42, 75) |
| Abbreviations: BMI, body mass index; VE, vaccine effectiveness; VHA, Veterans Health Affairs.<br><sup>a</sup>See Supplemental Table 1 for definitions of variables in this table.<br><sup>b</sup>The adjusted variables include the following: age, BMI, cancer, congestive heart failure, chronic kidney disease, diabetes mellitus, hypertension, immunocompromised, VA priority level, race, sex, rurality. |  |  |  |  |

